# Assessing the implications of access to toilet and water facilities on the health of households in the Sunyani Municipality, Ghana

**DOI:** 10.1101/2023.12.22.23300463

**Authors:** Prince Philip Ankapong Asare, Antwi Joseph Barimah, Catherine Mensah, Leilat Iddris Munkaila, Henry Ofosu Addo

**Author notes:** Corresponding author: Henry Ofosu Addo.

## Abstract

**Background:** Uncontaminated water and adequate sanitation facilities are of major importance for limiting the incidence of infectious diseases. In Ghana, about 80% of people still do not have access to proper sanitation, including latrines, which have dramatic consequences on human health, dignity, security and the environment. Potable water coverage in the Sunyani municipality stands at 47 percent in the urban areas and 33.5 percent in the rural areas. Many of the households in the Sunyani Municipality do not have toilet facilities, putting pressure on the few existing public toilet facilities available.

**Methods:** Using a simple random sampling technique, a total of 500 households were selected for the study. Structured questionnaires were used to collect quantitative data. In addition, a hand-held global positioning system (GPS) receiver was used to pick geographic coordinates of various water and toilet facilities. Quantitative data were analyzed using SPSS version 25.

**Results:** The results indicate inconsistencies in the spatial distribution of toilet and water facilities with an average distance of 33 meters. The study further revealed the current system of public toilets operating in the towns cannot be resource intensive to meet households’ targets because they do not satisfy sanitation needs (p<0.001). On the contrary, it tends to rather create even more problems, thereby encouraging open defecation. Also, their impact on human and environmental health needs to be taken into account.

**Conclusion:** Access to potable water and improved toilet facility remains a challenge as most households do not have toilets within their homes. Financial constraints, distance travelled and poor condition of public toilets were the main factors determining utilization of public toilet facilities. The types of toilet facilities used in the Municipality influence disease prevalence. The prevalence of cholera, diarrhoea, typhoid, skin rashes and eye infections were as a result of improper or no washing of hands.

## Introduction

Access to adequate sanitation and clean water is fundamental for societal progress, serving as a benchmark for civilization in modern society. However, despite global initiatives, significant portions of the global population still lack improved sanitation and access to clean water sources, perpetuating health risks and impeding societal advancement (1–3). An estimated 2.4 billion people still lack access to improved sanitation and 946 million still practice open defecation (4). It is estimated that 2.7 billion people use on-site sanitation worldwide and 1.77 billion use some kind of a pit latrine (5). Of those using some form of pit latrines, 65% of them are found in Sub-Saharan Africa (6). With access to potable drinking water globally, approximately 2 billion people continue to drink water contaminated with faeces (7).

Improved sanitation is measured by toilets with water-based architecture that flush into sewers and other septic systems, or pit latrines which can be simple pit latrines or pit latrines with improved ventilation (8). Latrines are excreta disposal facilities that can safely separate human excreta from human and insect contact (9).

Diseases attributable to inadequate water, sanitation and hygiene account for more than 4% of all disease burdens and deaths (4,10). Evidence from literature strongly shows the reduction in diarrhoea morbidity and subsequent mortality with such improved sanitation (11,12).

The relationship between water, sanitation, hygiene and public health has gained much prominence within public health circles across the globe. Per epidemiological and medical studies, the lack of sanitary toilets which is a key feature in developing countries exacerbates the transmission of bacteria especially *Salmonella* and *Escherichia coli* which invariably increases the onset of worms, malaria and other diarrhoeal infections (5).

Within the context of Ghana, substantial challenges persist in ensuring widespread access to safe water and improved sanitation. A majority of Ghanaians do not have access to toilet facilities in their homes and compounds (13). Open defecation is reported to be practiced by 15.2% of Ghanaian households (14). The availability and usage of public toilets remain the only alternatives to open defecation for a significant number of people in many low-income, urban communities (15). Ghana remains the country with the highest reliance on shared sanitation facilities globally (15).

Conspicuously, the Sunyani Municipality in the Bono region grapples with alarming deficiencies in these vital facilities, posing severe health risks to its residents (16). High rates of households lacking access to adequate toilets within their homes or compounds, coupled with prevalent open defecation practices, amplify health threats, particularly affecting vulnerable populations, such as children under five years old (16).

Despite Sunyani’s burgeoning urbanization, a noticeable gap exists in the literature regarding the direct relationship between inadequate access to improved toilet facilities and clean water sources and the resultant health implications for residents.

The Sunyani Municipality faces a pressing concern of inadequate access to improved toilet facilities and safe water sources, which significantly impact the health and well-being of its residents. The absence of detailed studies examining the direct correlations between these facilities and health outcomes within this rapidly growing urban area underscores the urgency of this research. Understanding the precise implications of inadequate sanitation and unsafe water on household health is essential for designing targeted interventions and policy frameworks aimed at ameliorating these challenges and ensuring a healthier living environment for Sunyani’s population. This research aims to address this critical gap by comprehensively assessing the multifaceted implications of water and toilet facilities on household health within the Sunyani municipality of Ghana.

In Ghana, 12.7% of households drink unsafe water and 80.6% use unimproved toilet facilities, with 18.8% resorting to open defecation (Ghana Statistical Service (GSS), Ghana Health Service (GHS) and ICF, 2018). A majority of Ghanaians do not have access to toilet facilities in their homes and compounds (13). According to Armah-Attoh (2015), approximately 9% of Ghanaians say they do not have access to toilet facilities. Availability and access to potable drinking water and improved sanitation are therefore of utmost significance to health and well-being (14). Open defecation is reported to be practiced by 15.2% of Ghanaian households (14). The availability and usage of public toilets remain the only alternatives to open defecation for a significant number of people in many low-income, urban communities (15). Ghana remains the country with the highest reliance on shared sanitation facilities globally (15).

Diarrhoeal diseases emanating from drinking contaminated water is one of the most common illnesses reported from health facilities in Ghana (14). Approximately, a quarter of all mortalities recorded among under 5 year old children in Ghana are attributable to diarrhoea (18). Sunyani, located in the then Brong Ahafo region has been reported to be one of the fastest growing cities in Ghana (16). According to the Afrobarometer data, the proportion of the population without access to toilets inside their homes or compounds in the Brong Ahafo region stood at 80% (13). This is relatively higher. The prevalence of open defecation among households in the Brong Ahafo region is estimated as 14.2% (14). As with any growing city in most developing countries, Sunyani municipality has its fair share of inadequate access to potable water and improved sanitation and its implication on health. There is a paucity of published work, however, on the implication to access to improved toilet facilities and potable water on the health of residents in the Sunyani municipality. This therefore, necessitated the need to assess the implications of water and toilet facilities on the health of households within the Sunyani municipality of Ghana.

## Methodology

### Study Area and design

The study was conducted in the Sunyani municipality of the Bono Region of Ghana. It lies between Latitudes 20°N and 70.05°N and Longitudes 20.30°W and 20.10°W. The study was descriptive cross-sectional in nature which explored the impact of water and toilet facilities on the health of households in the municipality. The study used Global Positioning System (GPS) device to collect coordinates of the water and toilet facilities in the municipality. The study population was composed of households and individuals within the Sunyani Municipality.

### Sampling

In this study, the sampling frame consisted of households in the Sunyani Municipality. A total of 1000 individual households were randomly selected to participate in the study, therefore the researchers used intuitive method i.e. 50% of the sampling frame to arrive at the required sample size of 500 respondents (Table 1).

**Table 1:** Sample Size calculation.

| Towns | Sample Size | Percentage % | Sampling frame |
| --- | --- | --- | --- |
| Abesim | 75 | 15 | 150 |
| Atronie | 100 | 20 | 200 |
| Sunyani Zongo | 50 | 20 | 200 |
| Benue Nkwanta | 50 | 10 | 100 |
| Nkrankrom | 25 | 5 | 50 |
| Wawasua | 75 | 15 | 150 |
| Atuahenekrom | 50 | 10 | 100 |
| Antwikrom | 25 | 5 | 50 |
| Bakoniaba | 50 | 10 | 100 |
| Total | 500 | 100 | 1000 |

### Data Collection Tools and techniques

#### Primary Data (Field)

The information from primary source was more reliable since it was gathered from questionnaires administered solely for the purpose of the study and the use of GPS device to collect coordinates of the water and toilet facilities to measure whether or not there are significant relationships between the distance of facilities (water and toilet) and the health of the households. In order to collect reliable and valid information, the researchers contacted some Departments within the Sunyani Municipal Assembly like the Community Water and Sanitation Agency. The primary data were collected between March and June 2022.

### Instrumentation

The questionnaires were distributed to the sampled respondents to know how they comprehended the implication of water and toilet facilities on the health of households, its sustainability, the challenges associated with it and the way forward. The researchers adopted a number of methods to get the required information for the study which included administration of standardized questionnaire, observation and the use of a hand-held Global Positioning System (GPS) device to map the coordinates of toilet and water facilities within the municipality.

### Study variables

The key variables in the study include the types of water and toilet facilities that are used in the Sunyani Municipality; the distance of the water and toilet facilities using the GPS device and the cost for accessibility and the health-related issues associated with the facilities and its usage.

### Data Processing and Analysis

Descriptive statistical tools such as percentages, bar graphs and cross-tabulations were used to present data collected in summarized charts and graphical forms where necessary to enhance visual appreciation of data collected using SPSS version 25.0. Also, a statistical test was used to determine the relationship between variables through the use of t-test that was used to determine relationship between independent variables and dependent variables. In this study, the statistical significance, dependence level was chosen to be 5% (i.e. p-value < 0.05). Further analysis was conducted using the ArcGIS version 10.4.1, GIS software to plot the coordinates in order to give a visual representation for the collected data.

### Ethical Considerations

Appropriate clearance was sought from requisite institutional review board prior to the commencement of the study. Informed consent was obtained from study participants and they were assured of utmost privacy and confidentiality.

## Results

### Presentation of Results

The study consisted of 500 respondents, out of these, 62% were females (Table 2). Sixty percent (60%) had a household size of 4-6, 34% were in a household size of 1-3 and 2% were in the largest household size of 11-13. With regards to the level of education, 30% had no formal education, 21% were primary school leavers, 20% were JHS/Middle school leavers and 9% had attained tertiary education (Table 2).

**Table 2:** Background attributes of respondents (N=500)

| **Variables** | **Frequency** | **Percentage (%)** |
| --- | --- | --- |
| **Gender** |  |  |
| Male | 189 | 37.8 |
| Female | 311 | 62.2 |
| **Age** |  |  |
| 18-29 | 119 | 23.8 |
| 30-39 | 160 | 32.0 |
| 40-49 | 101 | 20.2 |
| 50-60 | 70 | 14.0 |
| 70+ | 50 | 10.0 |
| **Marital status** |  |  |
| Single | 249 | 49.8 |
| Married | 225 | 45.0 |
| Divorced | 16 | 3.2 |
| Others | 10 | 2.0 |
| **Household size** |  |  |
| 1-3 | 171 | 34.2 |
| 4-6 | 302 | 60.4 |
| 7-10 | 17 | 3.4 |
| 11-13 | 10 | 2.0 |
| **Educational level** |  |  |
| No education | 152 | 30.4 |
| Primary | 104 | 20.8 |
| JHS/Middle School | 101 | 20.2 |
| SHS | 98 | 19.6 |
| Tertiary | 45 | 9.0 |
| **Occupation** |  |  |
| Formal Employee | 82 | 16.4 |
| Business | 121 | 24.2 |
| Trader | 98 | 19.6 |
| Farmer | 160 | 32.0 |
| No employment | 39 | 7.80 |
| **Employment duration** |  |  |
| 1-5 | 146 | 29.2 |
| 6-10 | 90 | 18.0 |
| 11-15 | 120 | 24.0 |
| 16-20 | 105 | 21.0 |
| Others | 39 | 7.8 |

A chi-square test was conducted to test for an association between background attributes and awareness of health implications of water and toilet facilities. The results indicated a statistical relationship between sex (p<0.035), age (p<0.001), marital status (p<0.002), household size (p<0.012), educational level (p<0.001), occupation (p<0.012) and awareness of health implications of water and toilet facilities (Table 3).

**Table 3:**
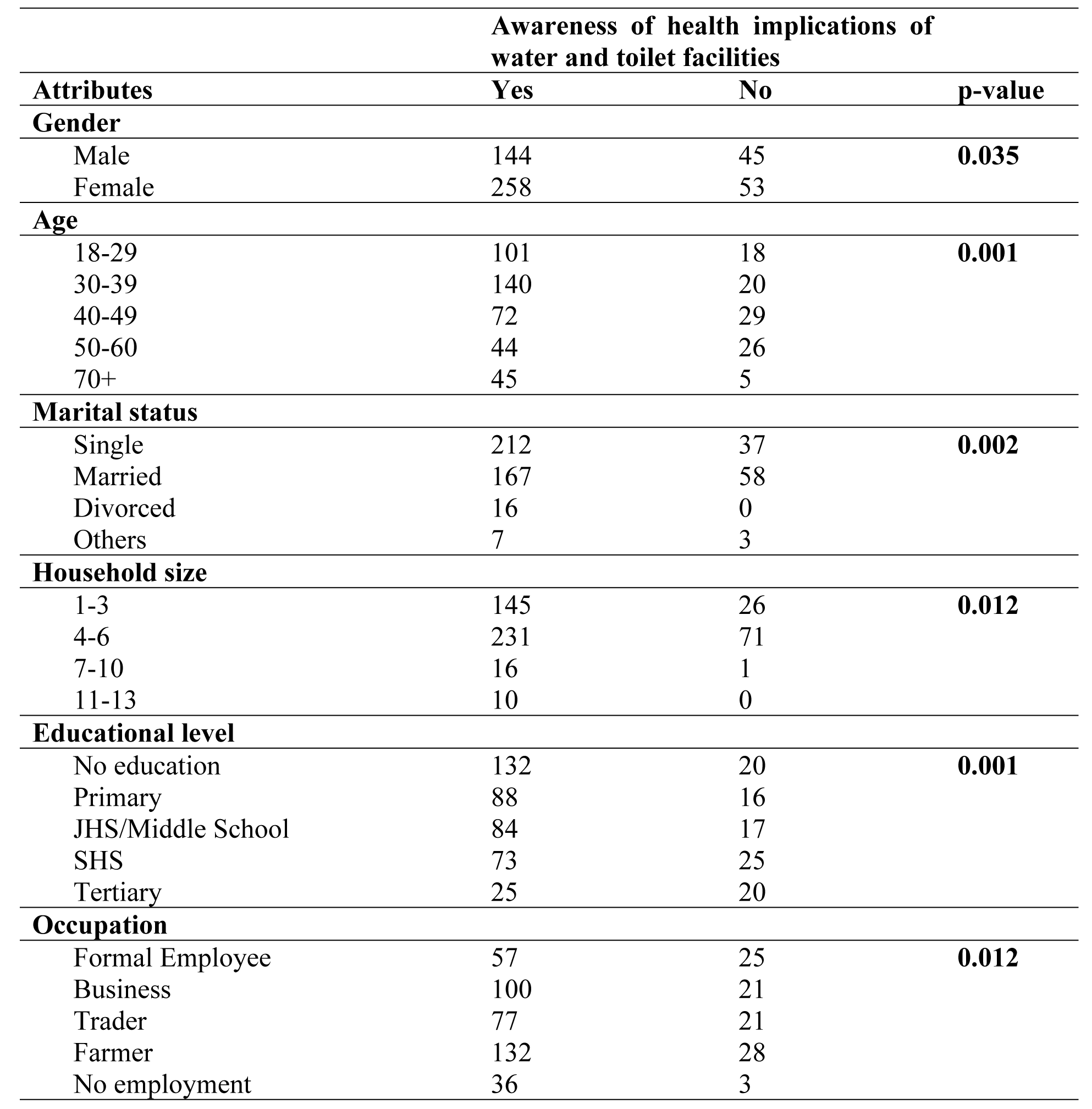
Relationship between background attributes and awareness of the health implications of water and toilet facilities.

|  |  | Awareness of health implications of water and toilet facilities |  |  |
| --- | --- | --- | --- | --- |
| Attributes |  | Yes | No | p-value |
| **Gender** |  |  |  |  |
| Male |  | 144 | 45 | **0.035** |
| Female |  | 258 | 53 |  |
| **Age** |  |  |  |  |
| 18-29 |  | 101 | 18 | **0.001** |
| 30-39 |  | 140 | 20 |  |
| 40-49 |  | 72 | 29 |  |
| 50-60 |  | 44 | 26 |  |
| 70+ |  | 45 | 5 |  |
| **Marital status** |  |  |  |  |
| Single |  | 212 | 37 | **0.002** |
| Married |  | 167 | 58 |  |
| Divorced |  | 16 | 0 |  |
| Others |  | 7 | 3 |  |
| **Household size** |  |  |  |  |
| 1-3 |  | 145 | 26 | **0.012** |
| 4-6 |  | 231 | 71 |  |
| 7-10 |  | 16 | 1 |  |
| 11-13 |  | 10 | 0 |  |
| **Educational level** |  |  |  |  |
| No education |  | 132 | 20 | **0.001** |
| Primary |  | 88 | 16 |  |
| JHS/Middle School |  | 84 | 17 |  |
| SHS |  | 73 | 25 |  |
| Tertiary |  | 25 | 20 |  |
| **Occupation** |  |  |  |  |
| Formal Employee |  | 57 | 25 | **0.012** |
| Business |  | 100 | 21 |  |
| Trader |  | 77 | 21 |  |
| Farmer |  | 132 | 28 |  |
| No employment |  | 36 | 3 |  |

In order to control for confounders and determine the predictors of respondents’ awareness of the health implications of water and toilet facilities in the Sunyani Municipality, a multiple logistic regression model was used (Table 4). The model took into consideration all significant variables at the simple logistic regression level, using an alpha value of 0.05. The result indicates that, females were less likely to have an awareness of health implications of water and toilet facilities. Respondents between the ages of 40-49 were ten times (OR 10.850, CI: 2.136-55.119) more likely to have an awareness of water and toilet facilities, those within 18-29 years (OR 0.780, CI: 0.344- 1.768) were less likely to have an awareness of the health implications of water and toilet facilities. It was also found that, JHS/Middle School leavers were more four times (OR 4.624, CI: 1.416- 15.094) more likely to have an awareness, SHS school leavers also six times (OR 6.025, CI: 1.979- 18.343) more likely to have awareness while those who had attained tertiary education were twenty-five times (OR 25.472, CI: 6.151-105.485) more likely to have an awareness. On occupation, traders were five times (OR 5.585, CI: 1.781-17.519) likely to have an awareness and those who were unemployed were less likely (OR 0.05, CI: 0.021-0.524) to have an awareness (Table 4).

**Table 4:** Background attributes and Awareness of health implications of water and toilet facilities (Multiple logistic regression)

| Background Determinants | Awareness of health implications of water and toilet facilities |  |  |
| --- | --- | --- | --- |
|  | Adjusted Odds Ratio | 95% CI |  |
| **Sex** |  |  |  |
| Male | **Ref** |  |  |
| Female | 0.137 | 0.059 | 0.316 |
| **Age (years)** |  |  |  |
| 18-29 | **Ref** |  |  |
| 30-39 | 0.780 | 0.344 | 1.768 |
| 40-49 | 10.850 | 2.136 | 55.119 |
| 50-60 | 4.205 | 0.874 | 20.236 |
| 70+ | 0.759 | 0.140 | 4.126 |
| **Educational level** |  |  |  |
| No education | **Ref** |  |  |
| Primary | 1.318 | 0.579 | 2.999 |
| JHS/Middle School | 4.624 | 1.416 | 15.094 |
| SHS | 6.025 | 1.979 | 18.343 |
| Tertiary | 25.472 | 6.151 | 105.485 |
| **Occupation** |  |  |  |
| Formal Employee | **Ref** |  |  |
| Business | 0.758 | 0.262 | 2.194 |
| Trader | 5.585 | 1.781 | 17.519 |
| Farmer | 0.627 | 0.206 | 1.908 |
| No employment | 0.105 | 0.021 | 0.524 |

### Types of water that exist in Sunyani Municipality

Table 5 describes the types of water sources that exist in the Municipality. Thirty-seven percent (37%) of respondents indicated borehole as their main source of water in the community, 20% relied on well/ponds, another 20% relied on pipe borne water and 12% relied on rivers as their source of water in the community. On the sources of water in the various households, 45% relied on boreholes, 21% on well/ponds; another 21% relied on pipe borne water and 5% on tanker service. Out of 100%, respondents (25%) indicated government or municipal assembly as the provider of their water, 36% stated the NGOs as the provider of their water and 16% indicated their community. Most of the respondents (70%) have their source of water outside their house, 20% had their source inside their house and 10% was outside the community (Table 5). Majority of the respondents (72%) did not treat their water before use while 28% treated their water. On the method used for water treatment, 13% boil their water, 3% use filters, 2% strain it through a filter and 1% allow it to settle (Table 5).

**Table 5:** Types of water (N=500)

| **Attributes** | **Frequency** | **Percentage (%)** |
| --- | --- | --- |
| **Main source of water in the community** |  |  |
| Borehole | 183 | 36.6 |
| Well/Ponds | 98 | 19.6 |
| River | 60 | 12.0 |
| Pipe Borne water | 102 | 20.4 |
| Rain water | 20 | 4.0 |
| Tanker service | 27 | 5.4 |
| Others | 10 | 2.0 |
| **Source of water in household** |  |  |
| Borehole | 228 | 45.6 |
| Well/Ponds | 106 | 21.2 |
| River | 25 | 5.0 |
| Pipe borne water | 106 | 21.2 |
| Rain water | 8 | 1.6 |
| Tanker service | 27 | 5.4 |
| **Who provides water used?** |  |  |
| Government/Municipal Assembly | 123 | 24.6 |
| NGO | 178 | 35.6 |
| Community | 81 | 16.2 |
| Household | 106 | 21.2 |
| Others | 12 | 2.4 |
| **Location of water source** |  |  |
| Inside house | 102 | 20.4 |
| Outside house (Community shared) | 349 | 69.8 |
| Outside community | 49 | 9.8 |
| **Treatment of water** |  |  |
| Yes | 140 | 28.0 |
| No | 360 | 72.0 |
| **Method of water treatment** |  |  |
| Boil | 67 | 13.4 |
| Strain it through cloth | 11 | 2.2 |
| Use a filter | 17 | 3.4 |
| Let it stand and settle | 6 | 1.2 |
| Others | 39 | 7.8 |
| None | 360 | 72.0 |

A bivariate analysis was conducted to ascertain the association between the outcome variable (awareness of the health implications of water and toilet facilities) and various independent variables (types of water). The results indicate that main sources of water in the community (p<0.001), the sources of water in the household (p<0.001) statistically influence respondents’ awareness of the health implications of water and toilet facilites (Table 6). It was also found that, location of water sources (p<0.002), treatment of water (p<0.002) and method of water treatment (p<0.003) were all statistically related to respondents’ awareness of the health implications of access to water and toilet facilities (Table 6).

**Table 6:** Relationship between types of water in the Municipality and awareness of the health implications of water and toilet facilities.

| Awareness of health implications of water and toilet facilities |  |  |  |
| --- | --- | --- | --- |
| Attributes | Yes: n (%) | No: n (%) | p-value |
| Main source of water in the community |  |  |  |
| Borehole | 178 (97.3) | 5 (2.7) | 0.001 |
| Well/Ponds | 66 (67.3) | 32 (32.7) |  |
| River | 35 (58.3) | 25 (41.7) |  |
| Pipe Borne water | 83 (81.4) | 19 (18.6) |  |
| Rain water | 17 (85) | 3 (15) |  |
| Tanker service | 20 (74.1) | 7 (25.9) |  |
| Others | 3 (30) | 7 (70) |  |
| Source of water in household |  |  |  |
| Borehole | 203 (89) | 25 (11) | 0.001 |
| Well/Ponds | 73 (68.9) | 33 (31.1) |  |
| River | 7 (28) | 18 (72) |  |
| Pipe borne water | 93 (87.7) | 13 (12.3) |  |
| Rain water | 6 (75) | 2 (25) |  |
| Tanker service | 20 (74.1) | 7 (25.9) |  |
| Who provides water used? |  |  |  |
| Government/Municipal Assembly | 110 (89.4) | 13 (10.6) | 0.002 |
| NGO | 173 (97.2) | 5 (2.8) |  |
| Community | 64 (79) | 17 (21) |  |
| Household | 51 (48.1) | 55 (51.9) |  |
| Others | 4 (33.3) | 8 (66.7) |  |
| Location of water source |  |  |  |
| Inside house | 84 (82.4) | 18 (17.6) | 0.001 |
| Outside house (Community shared) | 300 (86) | 49 (14) |  |
| Outside community | 18 (36.7) | 31 (63.3) |  |
| Treatment of water |  |  |  |
| Yes | 139 (99.3) | 1 (0.7) | 0.002 |
| No | 263 (73.1) | 97 (26.9) |  |
| Method of water treatment |  |  |  |
| Boil | 67 (100) | 0 (0) | 0.003 |
| Strain it through cloth | 11 (100) | 0 (0) |  |
| Use a filter | 17 (100) | 0 (0) |  |
| Let it stand and settle | 6 (100) | 0 (0) |  |
| Others | 38 (97.4) | 1 (2.6) |  |
| None | 263 (73.1) | 97 (26.9) |  |

From the multiple regression analysis, respondents who used borehole were three times (OR 3.671, CI: 2.046-6.585) more likely to be aware of the health implications of water and toilet facilities, those who used rain water were two times (OR 2.707, CI: 0.518-14.141) more likely to have an awareness as well as those who used tanker services (OR 2.842, CI: 1.093-7.391) (Table 7). Respondents who are provided with water by the community were three times (OR 3.991, CI: 1.317-12.094) more likely to have an awareness of health implications of water and toilet facilities. It was further revealed that respondents whose water sources are outside the community were more likely to have an awareness (OR 1.952, CI: 0.641-5.948) as well as those who treat their water before drinking (OR 0.021, CI: 0.003-0.141) (Table 7).

**Table 7:** Type of water used in Municipality and awareness of health implications of water and toilet facilities (Multiple logistic regression)

| **Determinants** | **Awareness of health implications of water and toilet facilities** |  |  |
| --- | --- | --- | --- |
|  | **Adjusted Odds Ratio** | **95% CI** |  |
| **Source of water in household** |  |  |  |
| Borehole | **Ref** |  |  |
| Well/Ponds | 3.671 | 2.046 | 6.585 |
| River | 20.880 | 7.940 | 54.906 |
| Pipe borne water | 1.135 | 0.556 | 2.317 |
| Rain water | 2.707 | 0.518 | 14.141 |
| Tanker service | 2.842 | 1.093 | 7.391 |
| **Who provides water used?** |  |  |  |
| Government/Municipal Assembly | **Ref** |  |  |
| NGO | 0.534 | 0.141 | 2.019 |
| Community | 3.991 | 1.317 | 12.094 |
| Household | 13.250 | 5.069 | 34.636 |
| Others | 7.845 | 1.531 | 40.200 |
| **Location of water source** |  |  |  |
| Inside house | **Ref** |  |  |
| Outside house (Community shared) | 0.414 | 0.156 | 1.100 |
| Outside community | 1.952 | 0.641 | 5.948 |
| **Treatment of water** |  |  |  |
| Yes | 0.020 | 0.003 | 0.141 |
| No | **Ref** |  |  |

### Types of Toilet Facilities that exist in the Municipality

From Table 8, 20% of respondents used private water closet in their community, 28% used public pit latrines, 6% used private pit latrine and 23% used other types of toilet facilities in the community. Fifty-eight percent (58%) of the respondents had their toilets located outside their house, 26% had it inside their house and 2% had their toilet facilities located outside their community. Out of 100%, 64% indicated the number of toilet facilities in the community as 1-3, 26% indicated 4-6 while 10% indicated that there are no toilet facilities in their community. With respect to those who have toilet facilities in their community, 43% are managed by the District Assembly, 19% are managed by private contractors and 28% are managed by area/urban council. Out of 90%, 84% were unsatisfied with the level of cleanliness of toilet facilities while only 6% showed satisfaction (Table 8).

**Table 8:**
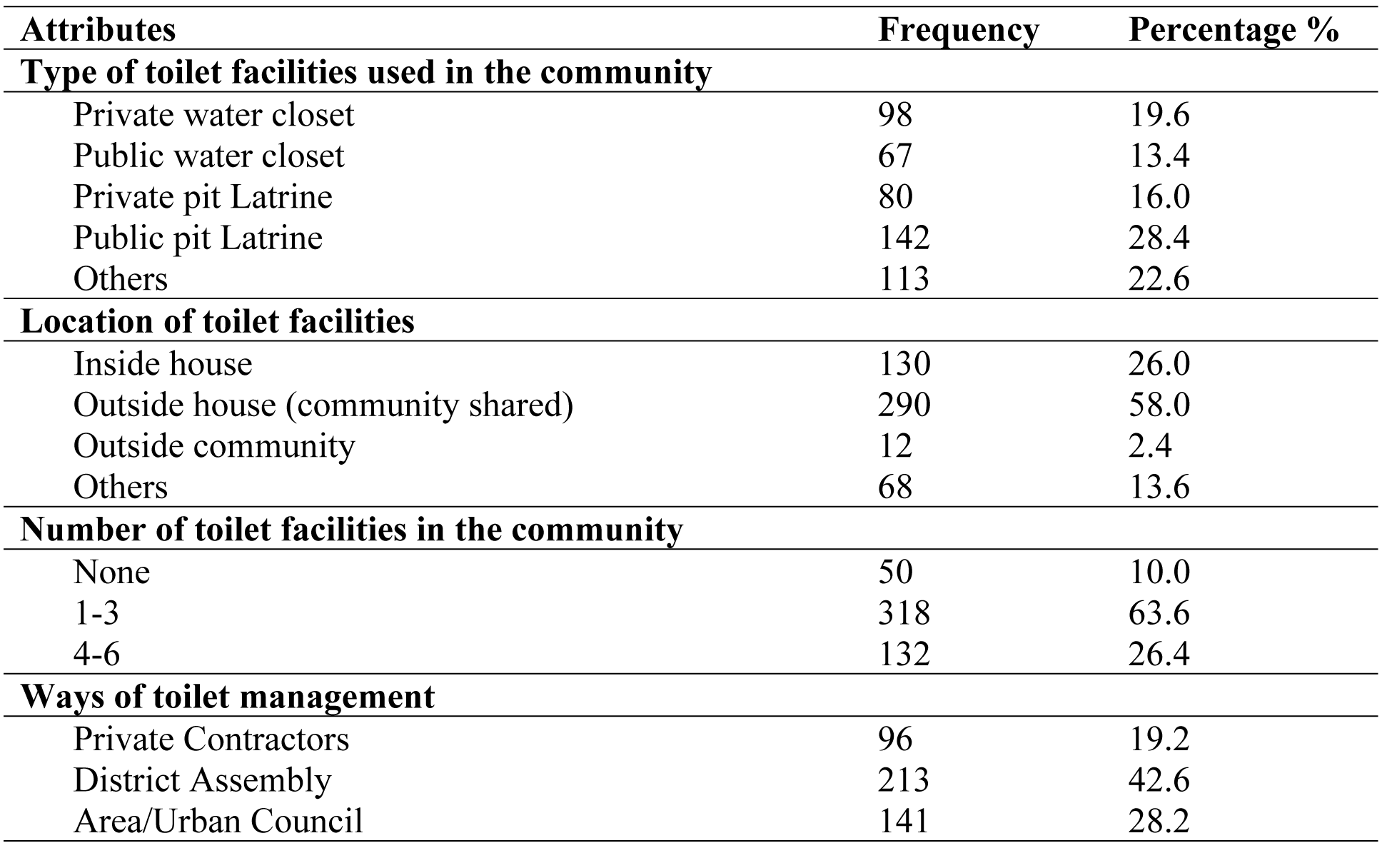
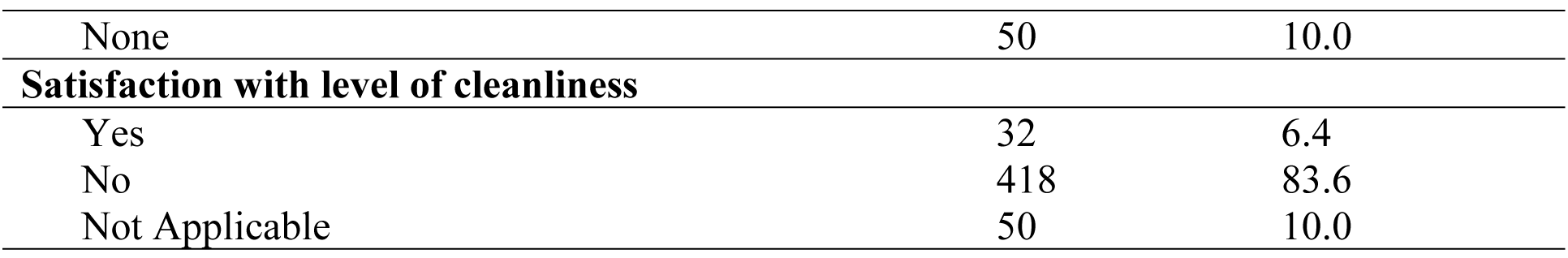
Types of Toilet Facilities (N=500)

| **Attributes** | **Frequency** | **Percentage %** |
| --- | --- | --- |
| **Type of toilet facilities used in the community** |  |  |
| Private water closet | 98 | 19.6 |
| Public water closet | 67 | 13.4 |
| Private pit Latrine | 80 | 16.0 |
| Public pit Latrine | 142 | 28.4 |
| Others | 113 | 22.6 |
| **Location of toilet facilities** |  |  |
| Inside house | 130 | 26.0 |
| Outside house (community shared) | 290 | 58.0 |
| Outside community | 12 | 2.4 |
| Others | 68 | 13.6 |
| **Number of toilet facilities in the community** |  |  |
| None | 50 | 10.0 |
| 1-3 | 318 | 63.6 |
| 4-6 | 132 | 26.4 |
| **Ways of toilet management** |  |  |
| Private Contractors | 96 | 19.2 |
| District Assembly | 213 | 42.6 |
| Area/Urban Council | 141 | 28.2 |
| None | 50 | 10.0 |
| **Satisfaction with level of cleanliness** |  |  |
| Yes | 32 | 6.4 |
| No | 418 | 83.6 |
| Not Applicable | 50 | 10.0 |

From the Table 9, the type of toilet facilities used in the community (p<0.001), the location of toilet facilities (p<0.010) and the number of toilet facilities in the community had a statistical relationship to respondents’ awareness of the health implications of water and toilet facilities. The study also revealed that, management of toilet facilities (p<0.003) and satisfaction with the level of cleanliness (p<0.001) influenced respondents’ awareness of the health implications of water and toilet facilities (Table 9).

**Table 9:** Relationship between types of toilet facilities in the Municipality and awareness of the health implications of water and toilet facilities.

| Awareness of health implications of water and toilet facilities |  |  |  |
| --- | --- | --- | --- |
| Attributes | Yes: n (%) | No: n (%) | p-value |
| Type of toilet facilities used in the community |  |  |  |
| Private water closet | 98 (100) | 0 (0) | 0.001 |
| Public water closet | 67 (100) | 0 (0) |  |
| Private pit latrine | 31 (38.8) | 49 (61.2) |  |
| Public pit latrine | 114 (80.3) | 28 (19.7) |  |
| Others | 92 (81.4) | 21 (18.6) |  |
| Location of toilet facilities |  |  |  |
| Inside house | 100 (76.9) | 30 (23.1) | 0.010 |
| Outside house (community shared) | 242 (83.4) | 48 (16.6) |  |
| Outside community | 12 (100) | 0 (0) |  |
| Others | 48 (70.6) | 98 (19.6) |  |
| **Number of toilet facilities in the community** |  |  |  |
| None | 0 (0) | 50 (100) | **0.001** |
| 1-3 | 270 (84.9) | 48 (15.1) |  |
| 4-6 | 132 (100) | 0 (0) |  |
| **Ways of toilet management** |  |  |  |
| Private Contractors | 96 (100) | 0 (0) | **0.003** |
| District Assembly | 165 (77.5) | 48 (22.5) |  |
| Area/Urban Council | 141 (100) | 0 (0) |  |
| None | 0 (0) | 50 (100) |  |
| **Satisfaction with level of cleanliness** |  |  |  |
| Yes | 12 (37.5) | 20 (62.5) | **0.001** |
| No | 390 (93.3) | 28 (6.7) |  |
| Not Applicable | 0 (0) | 50 (100) |  |

### Distance from water facilities to households and cost of accessibility

The study further assessed the distance from respondents’ houses to their water sources. It was found that, 39% travelled a distance of 101-200 meters to get their water, 21% travelled 50-100 meters, 20% travelled less than 50 meters, and 20% travelled over 201 meters to get water in their communities (Table 10). Eighty-one percent (81%) of respondents made no payment for their water use while 19% affirmed to payment for their water use. Out of those who made payment, 61% paid less than 50 pesewas and 20% paid 50-90 pesewas (Table 10). The maintenance of these water facilities is equally important in ensuring access, 69% indicated the water facility had broken down for 2-4 times in the past 12 months, 18% indicated once and 5% stated 5-10 times. Almost all of the respondents (99%) disagreed that water sources are available all year round. Out of these, 63% used borehole when their water sources break down, 31% resorted to wells/pond/spring and 6% resorted to other means (Table 10).

**Table 10:**
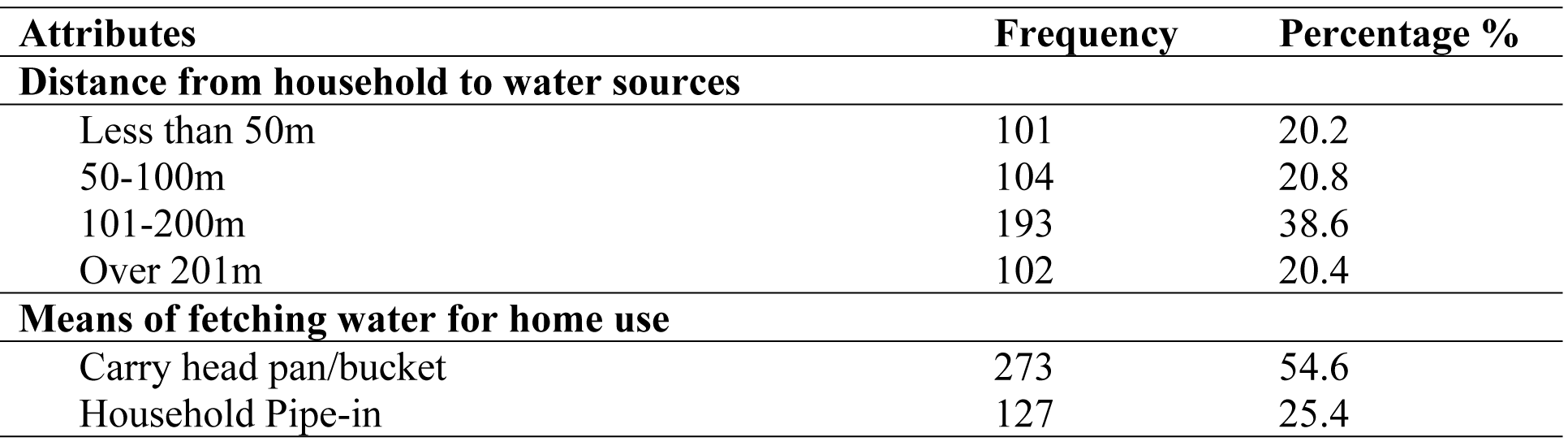
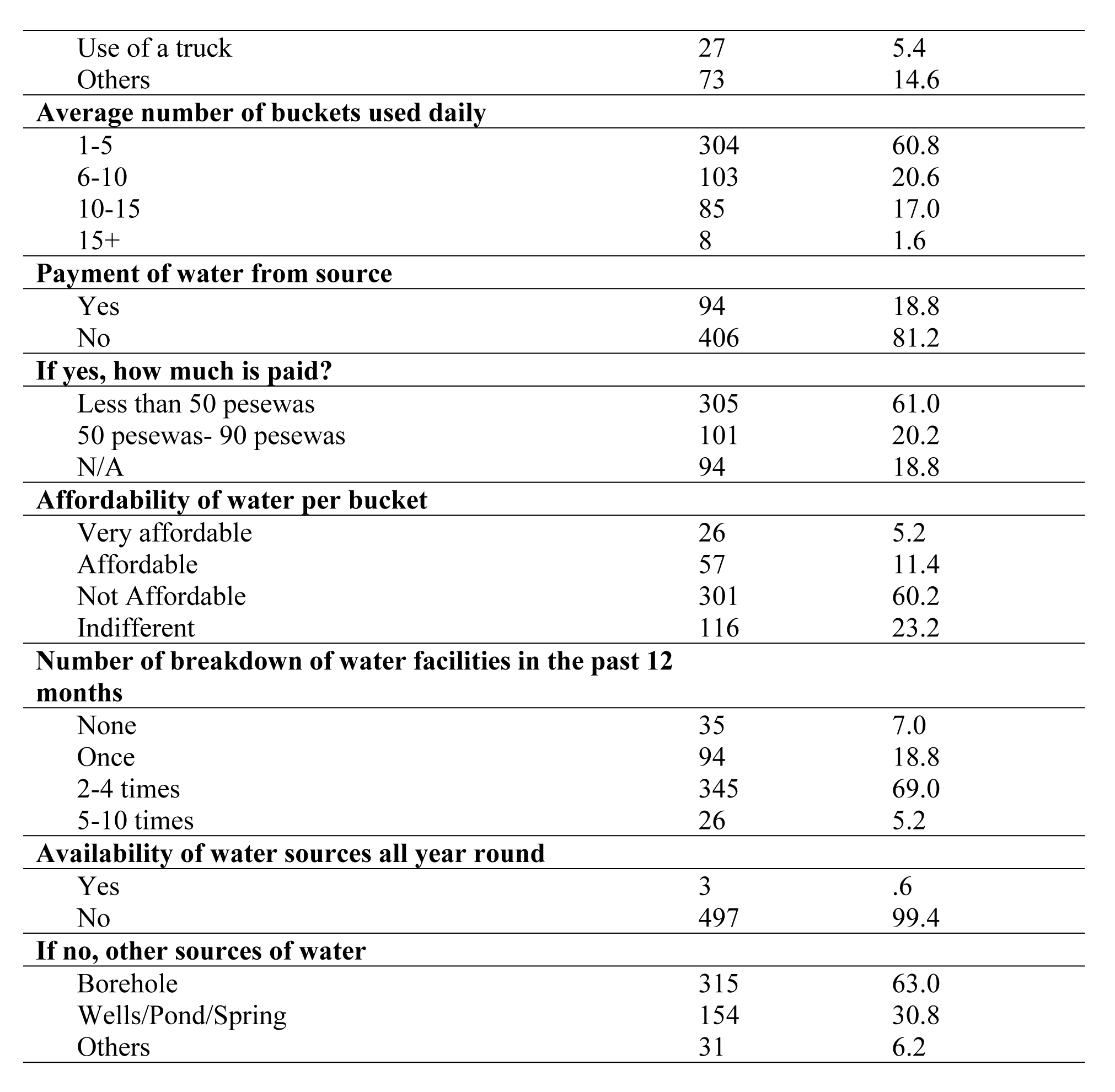
Distance from houses to water facilities and cost of accessibility (N=500)

| **Attributes** | **Frequency** | **Percentage %** |
| --- | --- | --- |
| **Distance from household to water sources** |  |  |
| Less than 50m | 101 | 20.2 |
| 50-100m | 104 | 20.8 |
| 101-200m | 193 | 38.6 |
| Over 201m | 102 | 20.4 |
| **Means of fetching water for home use** |  |  |
| Carry head pan/bucket | 273 | 54.6 |
| Household Pipe-in | 127 | 25.4 |
| Use of a truck | 27 | 5.4 |
| Others | 73 | 14.6 |
| **Average number of buckets used daily** |  |  |
| 1-5 | 304 | 60.8 |
| 6-10 | 103 | 20.6 |
| 10-15 | 85 | 17.0 |
| 15+ | 8 | 1.6 |
| **Payment of water from source** |  |  |
| Yes | 94 | 18.8 |
| No | 406 | 81.2 |
| **If yes, how much is paid?** |  |  |
| Less than 50 pesewas | 305 | 61.0 |
| 50 pesewas- 90 pesewas | 101 | 20.2 |
| N/A | 94 | 18.8 |
| **Affordability of water per bucket** |  |  |
| Very affordable | 26 | 5.2 |
| Affordable | 57 | 11.4 |
| Not Affordable | 301 | 60.2 |
| Indifferent | 116 | 23.2 |
| **Number of breakdown of water facilities in the past 12 months** |  |  |
| None | 35 | 7.0 |
| Once | 94 | 18.8 |
| 2-4 times | 345 | 69.0 |
| 5-10 times | 26 | 5.2 |
| **Availability of water sources all year round** |  |  |
| Yes | 3 | .6 |
| No | 497 | 99.4 |
| **If no, other sources of water** |  |  |
| Borehole | 315 | 63.0 |
| Wells/Pond/Spring | 154 | 30.8 |
| Others | 31 | 6.2 |

From the Table 11, 47% of the respondents travelled 5-10 minutes from their household to toilet facilities, 40% travelled less than 5 minutes and 13% travelled for 11-15 minutes to access toilet facilities (Table 11). Seventy-one percent (71%) agreed to payment for toilet facilities while 29% did not make payments (Table 11). Out of those who made payments, 59% paid 20-50 pesewas, 5% paid less than 20 pesewas and 6% paid 60 pesewas to 1 cedi. On its affordability, 54% indicated that the toilet facility was affordable, 12% indicated that it was very affordable and 21% were indifferent. Majority (86%) had no toilet facilities all year round and their alternative was “wrap and throw” by 84%, 14% used the ‘cat method’ and 2% used other methods (Table 11).

**Table 11:**
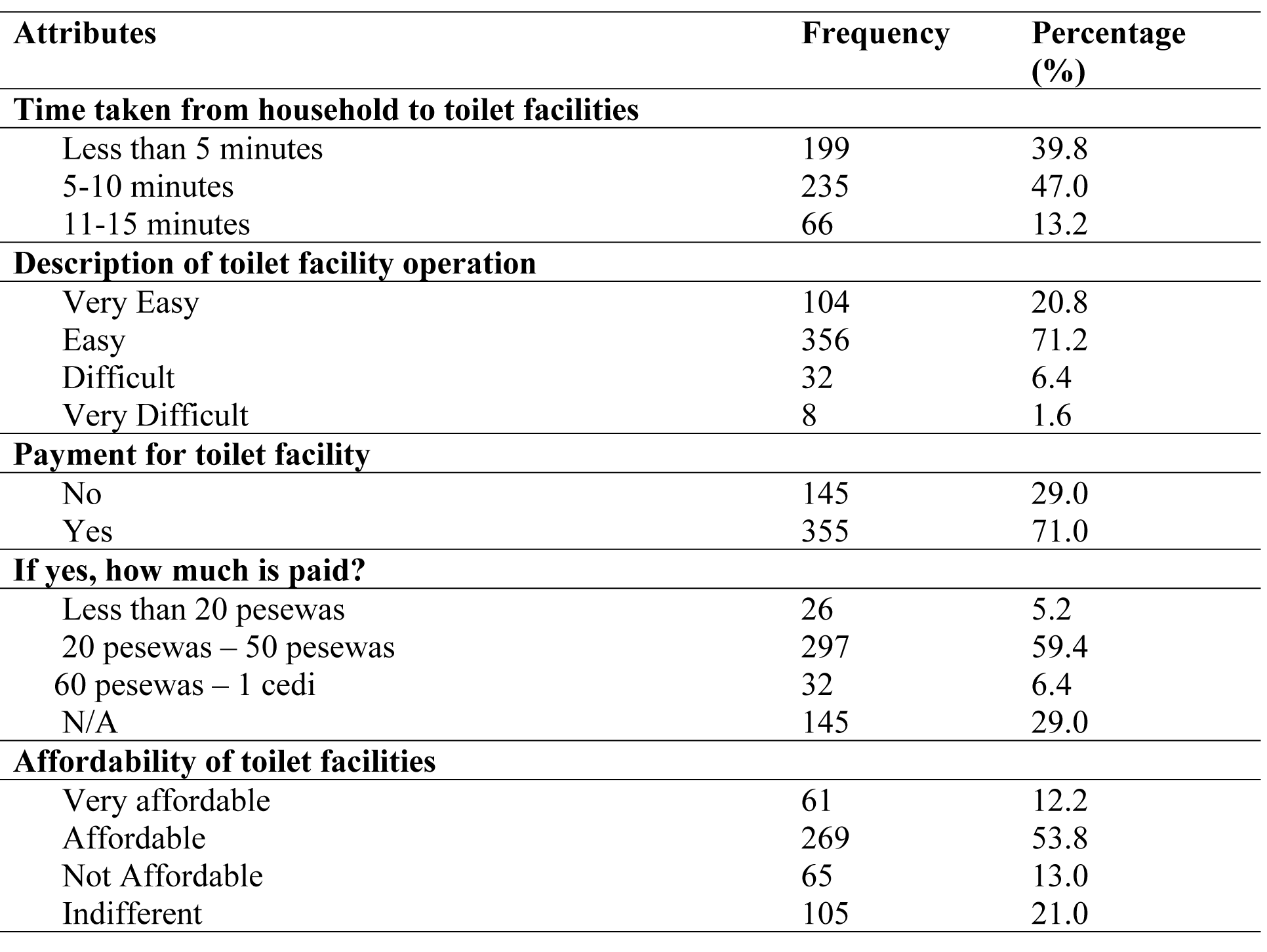
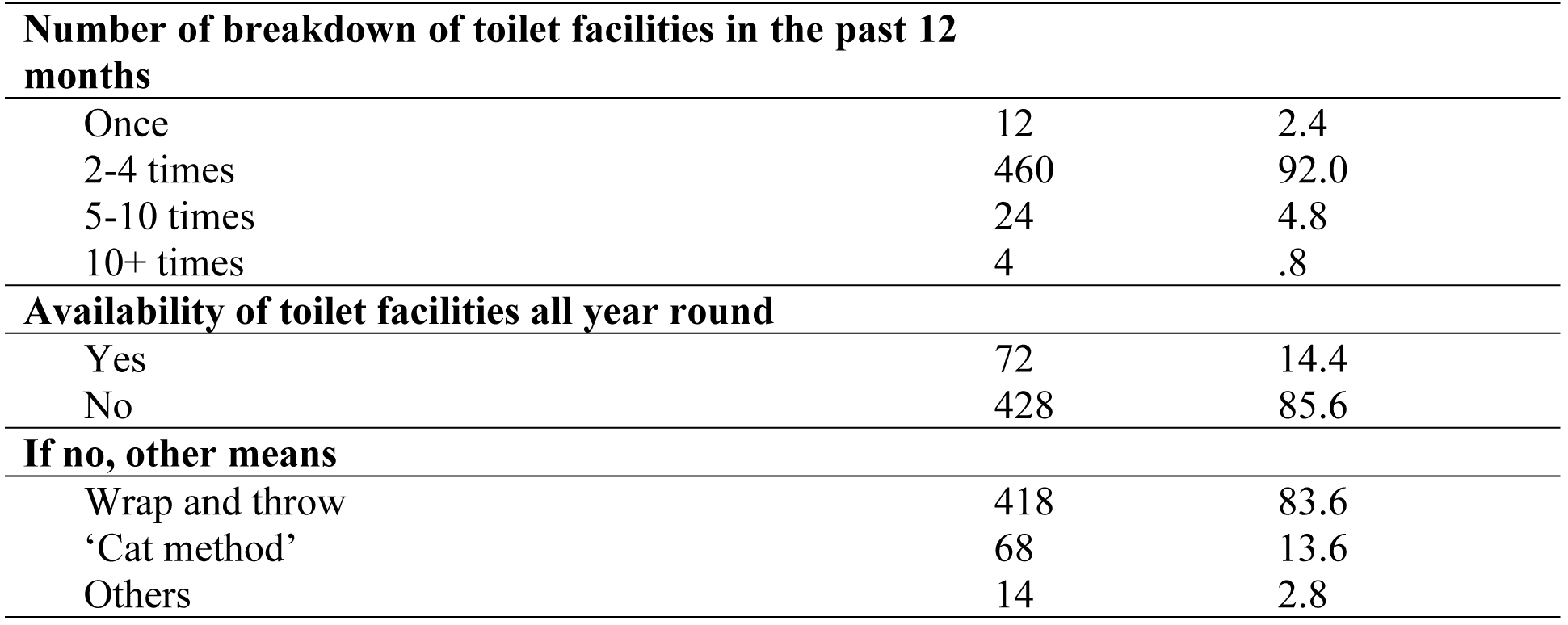
Distance from households to toilet facilities and cost of accessibility (N=500)

### Health related issues associated with water and toilet facilities usage

On the issues associated with water and toilet facilities usage, 33% used water for washing, 30% for drinking, 23% for cleaning and 13% for bathing (Table 12). Eighty-five percent (85%) of the respondents indicated they had developed health implications as a result of the use of poor water and toilet. Most of the respondents (75%) washed their hands after visiting the toilet, 25% used soap in hand washing, 49% used other methods and 26% did not practice hand washing (Table 12).

**Table 12:** Issues associated with water and toilet facilities usage.

| **Attributes** | **Frequency** | **Percentage (%)** |
| --- | --- | --- |
| **Uses of water in the household** |  |  |
| Cooking | 2 | .4 |
| Cleaning | 114 | 22.8 |
| Washing | 166 | 33.2 |
| Bathing | 68 | 13.6 |
| Drinking | 150 | 30.0 |
| **Use of water for income generation** |  |  |
| Yes | 125 | 25.0 |
| No | 375 | 75.0 |
| **Health implication as a result of poor water and toilet facility** |  |  |
| Yes | 426 | 85.2 |
| No | 74 | 14.8 |
| **Washing of hands after visiting toilet** |  |  |
| Yes | 376 | 75.2 |
| No | 124 | 24.8 |
| **What is used for hand washing?** |  |  |
| Soap | 125 | 25.0 |
| None | 131 | 26.2 |
| Others | 244 | 48.8 |

Table 13 represents diseases associated with water and toilet facilities usage and its frequency of occurrences. Majority of respondents (248) have had diarrhoea once in the last 12 months, 159 respondents have had diarrhoea 2-3 times and 2 respondents for 4-5 times in the last 12 months. Eighty-eight respondents had typhoid fever once in the last 12 months and 2 respondents for 2-3 times. Out of 500 respondents, only one reported of tuberculosis in the past 12 months while 174 respondents had cholera once in the last 12 months (Table 13).

**Table 13:** Diseases associated with water and toilet facilities usage and frequency of its occurrence.

| **Frequency of diseases in the last 12 months** | **Diarrhoea** |  |  |
| --- | --- | --- | --- |
|  | **Yes** | **No** | **Total** |
| Once | 248 | 1 | 249 |
| 2-3 times | 159 | 1 | 160 |
| 4-5 times | 2 | 0 | 2 |
| N/A | 0 | 89 | 89 |
| Total | 409 | 91 | 500 |
|  | **Typhoid fever** |  |  |
|  | **Yes** | **No** | **Total** |
| Once | 88 | 0 | 88 |
| 2-3 times | 2 | 0 | 2 |
| N/A | 55 | 355 | 410 |
| Total | 145 | 355 | 500 |

Table 13 represents diseases associated with water and toilet facilities usage and its frequency of occurrences.
|  | **Tuberculosis** |  |  |
| --- | --- | --- | --- |
| Once | 1 | 0 | 1 |
| N/A | 0 | 499 | 499 |
| Total | 1 | 499 | 500 |
|  | **Cholera** |  |  |
| Once | 174 | 0 | 174 |
| N/A | 0 | 326 | 326 |
| Total | 174 | 326 | 500 |

### Analysis on coordinates of toilet and water using GPS

As part of the research conducted to *“Assess the implication of water and toilet facilities on the health of households in the Sunyani Municipality, Ghana;* Global Positioning System (GPS) Coordinates were taken for each of the nine communities considered under this study; Atuahene, Atronie, Antwikrom, Wawasua, North Bosoma and the Sunyani town (Table 14). All private toilet, public toilet, public water and refuse dumps within each community were identified and further mapped using the GPS coordinates. Coordinates taken were further stored using the handheld GPS (GARMIN S10). **Table 14** shows coordinates of all facilities identified in each community.

**Table 14:** Coordinates obtained from various communities.

|  |  |  |
| --- | --- | --- |
| **Atuahene Community** |  |  |
| **Facility** | **N** | **W** |
| 944(pwuahii) | 7.11183 | -2.377639 |
| Private toilet | 7.111889 | -2.377917 |
| 946 | 7.111861 | -2.377917 |
| Public water | 7.1109722 | -2.3768056 |
| 948(pvti) | 7.1106944 | -2.377111 |
| 949(pt) | 7.112333 | -2.3760278 |
| 950(pvt) | 7.1120555 | -2.375694 |
| 951(pvt) | 7.112333 | -2.377083 |
| 952pw) | 7.1133056 | -2.378222 |
| **Atronie Community** |  |  |
| **Facility** | **N** | **W** |
| Pvt) | 7.152722 | -2.405694 |
| 954(pw) | 7.1523056 | -2.405778 |
| 955(pvt) | 7.1519167 | -2.405972 |
| 956(pw) | 7.1536389 | -2.4061667 |
| 957(pvt) | 7.1539444 | -2.406333 |
| 958(pt) | 7.1549722 | -2.4058056 |
| 959(rd) | 7.1549167 | -2.405889 |
| 960(pvt) | 7.1544167 | -2.4056389 |
| 961(pvt) | 7.1531667 | -2.4056944 |
| 962(pvt) | 7.15275 | -2.4048611 |
| **Antwikrom** |  |  |
| **Facility** | **N** | **W** |
| 963(antwikrom_pwi) | 7.2160556 | -2.3909167 |
| 964(pvt) | 7.2161389 | -2.3913056 |
| 965(pvt) | 7.215722 | -2.391222 |
| 966(bnpw) | 7.24889 | -2.364028 |
| 967(pt) | 7.248778 | -2.364306 |
| 968(pw) | 7.249528 | -2.364639 |
| 969(pvt) | 7.249472 | -2.364333 |
| 970(pw) | 7.2491111 | -2.362472 |
| 971(pt) | 7.249028 | -2.362028 |
| 972(pw) | 7.250583 | -2.362861 |
| **Wawasua Community** |  |  |
| **Facility** | **N** | **W** |
| 973(wawasua_pwi) | 7.28972 | -2.404583 |
| 974(pw) | 7.290417 | -2.406389 |
| 975(pt) | 7.2906111 | -2.406833 |
| 976(pvt) | 7.290972 | -2.404889 |
| 977(area_2pw) | 7.338028 | -2.322556 |
| 978(a2pt) | 7.338028 | -2.322528 |
| 979(dw) | 7.3381389 | -2.32269 |
| 980(pw) | 7.337944 | -2.3226667 |
| **North Bosoma** |  |  |
| **Facility** | **N** | **W** |
| 981(north_bosoma_pw) | 7.387333 | -2.3190278 |
| 982(pw) | 7.3369167 | -2.322278 |
| 983(pw) | 7.336833 | -2.3204167 |
| **Sunyani Town** |  |  |
| **Facility** | **N** | **W** |
| 984(sunyani_th) | 7.336722 | -2.32025 |
| 985(pw) | 7.33633 | -2.319139 |
| 986(pw) | 7.336111 | -2.318306 |
| 987(pw) | 7.335889 | -2.3183056 |
| 988(pt) | 7.335833 | -2.318194 |
| 989(pw) | 7.335806 | -2.3180556 |
| 990(pw) | 7.335833 | -2.31780556 |
| 991(pt) | 7.335444 | -2.3177222 |
| 992(pw) | 7.334139 | -2.3152 |
| 993(pt) | 7.334028 | -2.3151667 |
| 994(pt) | 7.334083 | -2.315194 |
| 995(pt) | 7.3342778 | -2.317528 |
| 996 | 7.3488056 | -2.342472 |

These coordinates were further imported into the Esri ArcGIS (Geographic Information System) version 10.4.1 environment for further analysis to generate a 2-dimentional map to give a visual representation to findings for easy understanding of all identified facilities and their proximity from each other.

Distances between facilities in each community were also measured using the ArcGIS software to determine their proximity from each other and the likely impact they may have on each other (Figure 1).

**Figure 1:**
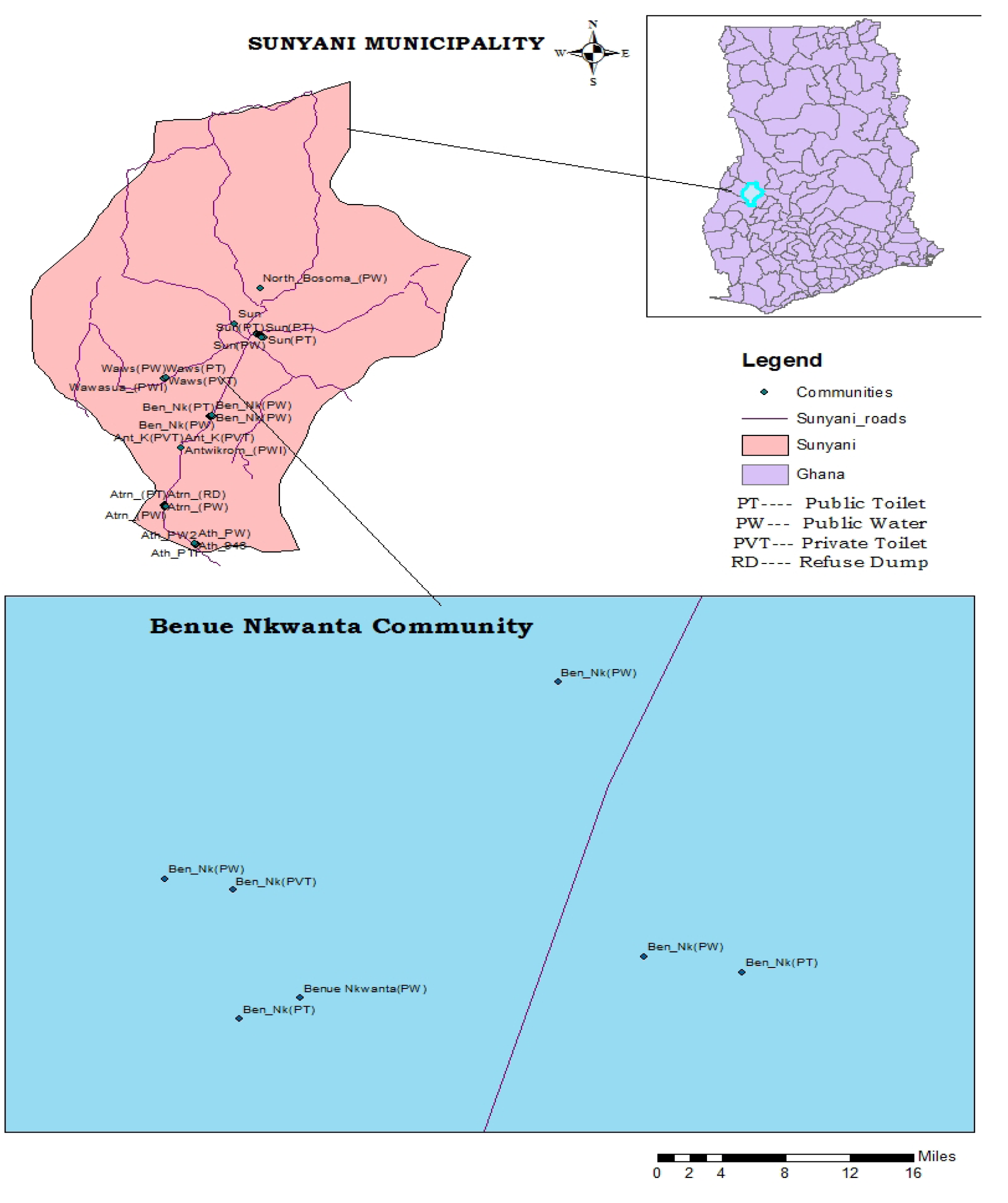
Map generated from coordinates

## Discussion

The provision of safe drinking water and improved toilet facilities still remains a challenge in most countries of the developing world (19,20). Globally, providing adequate sanitation is a challenge and the situation is worse in developing countries (6). Improved access to potable water and good sanitation protects the environment and improves public health.

The study found out that the main sources of water in the Sunyani Municipality and the households were boreholes and pipe borne water. However, boreholes were predominant over the pipe borne water from Ghana Water Company Limited (GWCL). The results indicated that there was significant relationship (p<0.001) between the type of water used in the household and the awareness of the health implication. This is corroborated by many studies which show the nexus between access to potable water and health (21,22). Even though respondents would prefer treated water from GWCL, it is only accessible to few and not readily accessible in remote areas leaving majority of the households relying on untreated water for domestic purposes. Respondents who used boreholes were more likely to have awareness of health implications of water and toilet facility whiles those who use rain water were less likely to have an awareness of health implications of water and toilet facilities.

The result indicated that majority of the respondents did not treat their water before use and the few who treated their water mostly did so by boiling. This study agrees with Addo et al. (2014), where most of the households they studied in three selected communities in Accra did not treat their water before use. In their study, Brown & Sobsey (2012) also reported that more than 90% of households they studied in Cambodia used boiling as a method of treating drinking water. The result further indicated relationship between treatment of water (p<0.002) and the method of treatment (p<0.003) to respondents’ awareness of the health implications of water and toilet facilities. Respondents who treat their water before use are more likely to have an awareness of health implications as opposed those who do not treat their water before use. Water treatment is very crucial for households who do not trust the source and quality of water they depend on. Hence, treatment of water before use becomes the best alternative (25). Asefa et al. (2023) also reported in their study from Southern Ethiopia that, 35.8% of their study participants practiced water treatment.

From this study, majority of the households who had to fetch water outside their households join long queues and as such spend longer time in accessing water. Women and children risk their lives in search of water, walking long distance in search of good water sources. The fetching of water can represent a substantial physical and economic burden that predominantly affects women and children (26). In most developing countries, fetching water for drinking and other household uses is a substantial burden that affects water quality and quantity in the household (20).

Water storage was noted as one of the major issues households battled with. This is expected due to inadequate education on water safety. The study revealed that majority of respondents collected water outside the house. It was revealed that the location of water sources (p<0.002) was statistically related to respondents’ awareness of the health implications of water and toilet facilities. Households contributed to the provision of water by way of monetary contribution and communal labour.

The types of toilet facilities in the Sunyani Municipality were public water closet, private water closet, public pit latrine and private pit latrine. This is consistent with the study of Kosoe & Osumanu (2018), where similar toilet facilities were found among respondents in Wa, Ghana. Majority of respondents relied on the public pit latrine. This study concurs with the study of Obeng et al. (2015), where as much as 47% of their respondents relied on pit latrines in a Ghanaian per- urban community. Regular use of latrines has been shown to be very useful to public health (28).

The result indicated that the type of toilet facilities used in the community (p<0.001), the location of toilet facilities (p<0.010) and the number of toilet facilities in the community had a statistical relationship to respondents’ awareness of the health implications of water and toilet facilities. The second largest type of toilet facility (22.6) was others (cat method, wrap and throw) due to inadequate toilet facilities in the municipality. The few toilet facilities which existed in the municipality were untidy leading to respondents’ high unsatisfactory level of cleanliness and thus low patronization. Several studies have also reported similar findings where patrons of public toilet facilities complained of the deplorable state of these toilet facilities (9,15,29). The study further revealed that, management of toilet facilities and satisfaction with the level of cleanliness influenced respondents’ awareness of the health implications of water and toilet facilities. The study revealed that majority of respondents visited toilet facilities outside the household. Again, the few that existed were poor and substandard toilet facilities.

In determining the distance of water source from the household, the study found that, majority of households (38.6%) had their water source within the distance of 101- 200 meters. The results also showed that, the minority of the households (20.2%) had their water sources at a distance less than 50 meters. This means that, the number of the households who walked longer distances to fetch water was higher than those who had their water access point closer. It therefore implies that, there is too much time and energy needed in fetching water and the level of contamination could be determined by the distance of water access point. Households with travel times more than 30 minutes have been shown to collect progressively less water (20).

It was also found that, majority (81.2%) of households do not pay for access to water and the few that pay (18.8%) claim they pay less than 50p, which they deem not affordable. The study again revealed that this high cost for accessibility was due to the increase in utility bills such as electricity. This study is not consistent with the study by Amoah (2020), where households spend more than GH₵84.30 ($14.70) on access to water. This is considered relatively higher.

Majority of households were within 5-10 minutes of toilet facilities. Many respondents (71.0%) made payment for toilet facility from a range of 20-50 pesewas which most (53.8%) deem affordable leading to majority of respondents (71.2%) describing the operations of the toilet facilities as easy. This agrees with the study by Arku et al. (2013) where respondents paid 10 and 20 pesewas to access toilet facilities. The cost of using a toilet facility ranged from 15 to 30 pesewas per use in low-income, Accra (15). Higher user fees to use public toilets, low level of education and unhygienic toilets have been identified as obstacles preventing access to sanitation facilities in urban slums in Ghana (31). The cost of public toilets was described as the most important reason for open defecation (15). In as much as an amount such as 20 Ghana pesewas may seem immaterial, it is quite expensive for individuals who do not have day jobs and even if they do, are being paid an amount unable to cater for their everyday use of toilet facilities. The expensive nature of these facilities has forced individuals moving out of the communities into open areas such as rivers, streams, farms openly defecating. Using the GPS device to map the coordinates of toilet and water facilities, it was realized that, the distance of public water to private toilet was 33.55 meters and the distance of private water to public toilet was 33.11meters. Distance to toilet facility is a key determinant of its usage. Arku et al. (2013) detailed in their study that respondents were not satisfied with having to walk over 500 metres before using a toilet facility. The odds of utilizing latrine in households with more than 6 metres latrine distance from households were 27.43 times higher than those who had less than or equal to 6 metres (32). According to the Sanitation Technologies in Emergencies (2023), the distance between households and shared toilet should be a maximum of 50 metres. Similarly, the distance between toilet and water sources at least 30 metres. This present study recorded a slightly higher distance of 33.11 metres between private water and toilet facility. In a study by Mabvouna et al. (2023), about 70.9% diarrhoea cases were observed in households with external latrines, and among them, 57.1% in households with latrines located at a distance less than 6 metres from households.

Majority of respondents (75.2%) washed their hands after using the toilet whiles 24.8% did not. This means that the 24.8% are highly susceptible of acquiring faeco-oral diseases and stand a chance of infecting the majority that wash their hands. Even though almost half of households in the municipality use public pit latrine, the majority that use other types of toilet facility stand a higher risk of diseases due to unhygienic conditions.

The study revealed four diseases associated with water and toilet facilities in the municipality in the last 12 months as diarrhoea, cholera, typhoid and tuberculosis in descending order. Majority of respondents 248 had diarrhoea once in the last 12 months while only one respondent reported of tuberculosis in the last 12 months.

## Conclusion

Inadequate access to clean water and sanitation is a major problem and is an integral part of Ghana’s economic development and poverty reduction policy. Despite the increased support provided to the sector, there are many people still depending on unsafe drinking water source, especially in the rural areas of the country. Those who collect the water (usually women and children) spend much of their time walking miles to carrying water. This leaves them little time for school, other work at home, or community life. Access to potable water and improved toilet facility remains a challenge as most households do not have toilets within their homes. Financial constraints, distance travelled and poor condition of public toilets were the main factors determining utilization of public toilet facilities. The study found that there was significant relationship (p<0.001) between the type of water and toilet used in the household, treatment of water (p<0.002) and the method of treatment (p<0.003), location of water sources and toilet facilities (p<0.002) and the awareness of the health implications of toilet and water facilities.

## Competing interest

The authors declared no potential conflicts of interest with respect to the research, authorship, and/or publication of this article.

## Sources of funding

The authors received no financial support for the research, authorship, and/or publication of this article.

## Authors’ contribution

HOA and PPAA conceptualized the overall study with its goals and aims. CM retrieved the requisite data from databases. AJB and LIM played a supportive role in data consolidation. AJB wrote the study background and played a supportive role in data analysis. HOA played a vital role in designing the methodology for the study. HOA and AJB played a role in writing the discussion of the study. PPAA, AJB, CM, LIM and HOA supported the writing, review, and editing of the manuscript. PPAA, CM and LIM did the data analysis for the study. All authors contributed significantly to the critical revision and approved the final version before the onward submission.

## Data Availability

All relevant data are within the manuscript and its Supporting Information files.

## Acknowledgement

Authors express their profound gratitude to respondents across the various households within the Sunyani municipality who consented to take part in the study.

## Notes

### Competing Interest Statement

The authors have declared no competing interest.

### Clinical Protocols

N/Ap

### Funding Statement

The author(s) received no specific funding for this work.

### Author Declarations

Ghana Health Service Ethics Review Committee with approval number GHS-ERC 509/02/22

